# Prospective cohort study of *Cryptosporidium* infection and shedding in infants and their households

**DOI:** 10.1101/2022.10.25.22281515

**Authors:** Poonum Korpe, Zhanmo Ni, Mamun Kabir, Masud Alam, Tahsin Ferdous, Rifat Ara, Rebecca M. Munday, Rashidul Haque, Priya Duggal

**Affiliations:** Department of Epidemiology, Johns Hopkins Bloomberg School of Public Health, Baltimore, MD; Emerging Infections and Parasitology Laboratory, International Centre for Diarrhoeal Disease Research, Bangladesh, Dhaka, Bangladesh; Department of Genetic Medicine, Johns Hopkins School of Medicine, Baltimore, Maryland, USA

**Keywords:** *Cryptosporidium*, Bangladesh, diarrheal disease

## Abstract

**Background:** *Cryptosporidium spp* are responsible for significant diarrheal morbidity and mortality in under-five children. There is no vaccine, thus a focus on prevention is paramount. Prior studies suggest that person-to-person spread may be an important pathway for transmission to young children. Here we describe a longitudinal cohort study of 100 families with infants to determine rates of cryptosporidiosis within households during the COVID-19 pandemic.

**Methods:** Families living in Mirpur, Bangladesh with one infant age 6-8 months were enrolled and followed with weekly illness survey and stool testing for Cryptosporidium for 8 months.

**Results:** From December 2020 to August 2021, 100 families were enrolled. Forty-four percent of index children, and 35% of siblings had at least one *Cryptosporidium* infection. Shedding of *Cryptosporidium* occurred for a mean of 19 days (sd 8.3 days) in index infants, 16.1 days (sd 11.6) in children 1-5 years, and 16.2 days (sd 12.8) in adults. A longer duration of *Cryptosporidium* shedding was associated with growth faltering in infants. There was a spike in *Cryptosporidium* cases in May 2021, which coincided with a spike in SARS-CoV-2 cases in the region.

**Conclusion:** In this intensive, longitudinal study of *Cryptosporidium* infection in families we found high rates of cryptosporidiosis in infants and children, and prolonged parasite shedding, especially among malnourished children. These data support that transmission within the household is an important route of exposure for young infants, and that treatment of non-diarrheal infection to interrupt person-to-person transmission within the home may be essential for preventing cryptosporidiosis in infants.

**summary:** Cryptosporidiosis is a leading cause of morbidity and mortality among children. We followed 100 families with infants living in Bangladesh and studied the incidence of *Cryptosporidium* infection. We found prolonged *Cryptosporidium* shedding in stool was common among infants and adults.

## BACKGROUND

*Cryptosporidium*, an enteric protozoa, has been identified as a leading contributor to moderate-to-severe diarrhea in children younger than 2 years of age worldwide [1]. Cryptosporidium diarrhea and subclinical infection have both been associated with stunted growth and cognitive deficits in children [2-5]. Despite the high morbidity from this infection, there is currently no effective treatment for infants younger than one year of age.

*Cryptosporidium* is transmitted by fecal-oral contamination. Transmission of diarrheal pathogens has been traditionally described as occurring along the “five-F pathways”: fluids, fingers, food, fields, and flies [6]. *Cryptosporidium* oocysts are readily infectious when excreted in feces, and ingestion of just 10 oocysts can result in infection [7]. *Cryptosporidium* are also resistant to chlorination [8,9]. Sporadic waterborne outbreaks have been well-documented in industrialized countries, but in endemic areas, like Bangladesh, infection is associated with poor sanitation, poverty, animal rearing, and malnutrition [3,10,11].

Efforts to combat *Cryptosporidium* infection include improving quality of drinking water and sanitation. However, non-pharmacologic studies to reduce environmental exposure and contamination have not been successful at preventing symptomatic or asymptomatic infections. In a semi-urban slum in India, where cryptosporidiosis is endemic, an intervention aimed at providing clean water to households found that children who drank bottled water did not have reduced rates of *Cryptosporidium* infection compared to those who drank from the municipal water supply [12]. Moreover, a community-based introduction of membrane filtered drinking water, with pore size small enough to filter *Cryptosporidium spp*, did produce microbiologically safe drinking water for 1 year, however, did not result in reduced rates of diarrhea in children under two [13]. The WASH Benefits study, a cluster randomized controlled trial, demonstrated success in providing clean water and improved handwashing and sanitation in communities in rural Bangladesh; however, despite these improved measures, there was no improvement in rate of cryptosporidiosis and there was no significant improvement in child growth [14]. This study highlighted that cryptosporidiosis may not be amenable to traditional WASH interventions because of its low infectious dose.

In a small, pilot study, we previously demonstrated that 39% of households with a Cryptosporidium-infected child had a second infected household member. Furthermore, genotyping of Cryptosporidia suggested that transmission in young children primarily occurs via human-to-human transmission rather than zoonosis and emphasized the importance of household transmission in *Cryptosporidium* infections for children under two years of age [15].

To further explore *Cryptosporidium* transmission, we enrolled a cohort of 100 families with an index child 6-8 months of age, and followed them prospectively for 8 months. The extensive longitudinal follow-up provides an opportunity to define incidence of *Cryptosporidium* infection within families with infants and to identify factors involved in transmission of the disease. Enrollment began during the early years of the COVID-19 pandemic, allowing a unique perspective on the effect of social distancing measures on transmission of an enteric protozoal infection.

## METHODS

### Study Design

Families with infants 6 to 8 months old living in Mirpur, Bangladesh were identified and approached for enrollment into the study. If families agreed to participate all household members, defined as any individual sleeping under the same roof or eating from the same cooking pot, were consented for enrollment. Once enrolled, a demographic enrollment form was completed for each participant. Baseline characteristics collected included age, weight, height, and familial relationships from all participants. At the household level, socioeconomic data including household income, parental occupation, maternal education, and crowding were collected. Additionally, data on water, sanitation, and household environment were collected, including source of drinking water, method of treatment of drinking water, toilet type, and presence of domesticated animals in the home.

Subsequently, all index infants and family members were surveyed weekly for any illness, including diarrhea and respiratory illness. A stool specimen was collected from each participant weekly, and during active diarrheal illness, over an 8-month follow up period. A serum specimen was collected from participants at baseline and at the of the 8-month follow up.

### Laboratory Testing

Stool and serum samples were collected in Mirpur and transported via cold chain to the Parasitology Laboratory at the International Centre for Diarrhoeal Disease Research, Bangladesh. Testing for *Cryptosporidium* was performed using the *Cryptosporidium* II ELISA kit (TechLab, Blacksburg, VA) on all weekly and diarrheal stool samples. SARS-CoV-2 IgG testing was performed using the TechLab SARS-CoV-2 ELISA (investigational use only) on baseline and 8-month serum samples collected from each participant.

### Case Definitions and Statistical Analysis

Symptoms were analyzed based on the weekly clinical survey of each participant. A *Cryptosporidium* case was defined as a participant having at least one stool sample positive for *Cryptosporidium* during the 8-month follow up period. A diarrheal episode was defined as 3 or more loose stools per day, recurring daily within a 14-day period. A diarrhea associated positive *Cryptosporidium* episode was defined as at least one positive diarrhea sample during the episode of infection.

The duration of *Cryptosporidium* shedding was calculated for each episode of *Cryptosporidium* infection. An episode was defined as stool samples testing positive within a 14-day period. The duration of the episode was calculated by the difference in days between the first positive stool sample in the episode and the first negative stool sample after the episode. If a positive sample was collected in the last week of study follow up, this was assigned a minimum 7-day duration of positivity.

The height-for-age adjusted z-scores and weight-for-age adjusted z-scores were calculated based on World Health Organization standards [16]. Lengths/heights and weights of the participants were measured monthly after baseline enrollment. The ages of children were rounded to the nearest month at time of each measurement.

#### Socioeconomic definitions

“Overcrowding” in the home was classified as >3 people per room per household [17]. Drinking water was categorized as water piped into the home, water piped into the yard, and tube well [18]. “Unimproved” toilets were categorized as having no facility or traditional pit latrine without slab. “Improved” toilet included ventilated improved pit toilet with water seal and flush toilet to piped sewer system, septic tank, or pit latrine [18].

#### Statistical analysis

Linear regression was used to estimate the difference in duration of shedding between index children vs other children vs adults, adjusting for sex and baseline COVID-19 IgG result. Linear regression was also applied to identify risk factors associated with longer duration of shedding. Both models are presented, and models with the lowest AIC based on stepwise model selection. Logistic regression was used to identify risk factors for *Cryptosporidium* positive cases at the household level. All tests were based on a two-sided p < 0.05. Fisher’s exact test was used for comparing group differences of categorical variables and Wilcoxon-Mann-Whitney test for continuous variables in exploratory analysis because of their relaxed assumptions on large sample sizes. Chi-square tests were used to compare difference in symptoms reported among weeks with *Cryptosporidium* infection and negative for infection among participants. Analysis was performed using R, version 4.2.0.

### Ethics Approval

The study was approved by the Research and Ethics Review Committees at the ICDDR,b and by the Institutional Review Board at the Johns Hopkins University Bloomberg School of Public Health. Written consent or assent was obtained from all participants over 11 years old, and parental consent was obtained for all children 11 years old and younger.

## RESULTS

From September 29, 2020 to December 22, 2020, field workers conducted a census in Mirpur Wards 2,3, and 5 and identified 1841 households. Of these, 284 families were identified as being eligible and were screened for enrollment. Between December 2020 and August 2021, 100 families were enrolled into the study, with a mean duration of follow up of 212.7 days (sd 28.8). Of the 100 families enrolled, 4 families were lost to follow up, and had a mean duration of follow up of 98.6 days (sd 54.2).

Families were comprised of 100 index children between ages 6-8 months, and 242 family members (Table 1, total n=342) including 100 mothers, 49 fathers and 60 siblings. Sociodemographic characteristics of this cohort were similar to prior studies in this area [3,15,19]. Seventeen percent of primary caregivers had no formal education. Overcrowding was found in 48% of households. Sixteen percent of households owned animals, and these were chickens or ducks. Nearly 1/3 of household had an unimproved toilet which includes no toilet or a pit latrine. Treatment of drinking water was common and included boiling water prior to drinking (64%) or using a filter (6%).

**Table 1.**
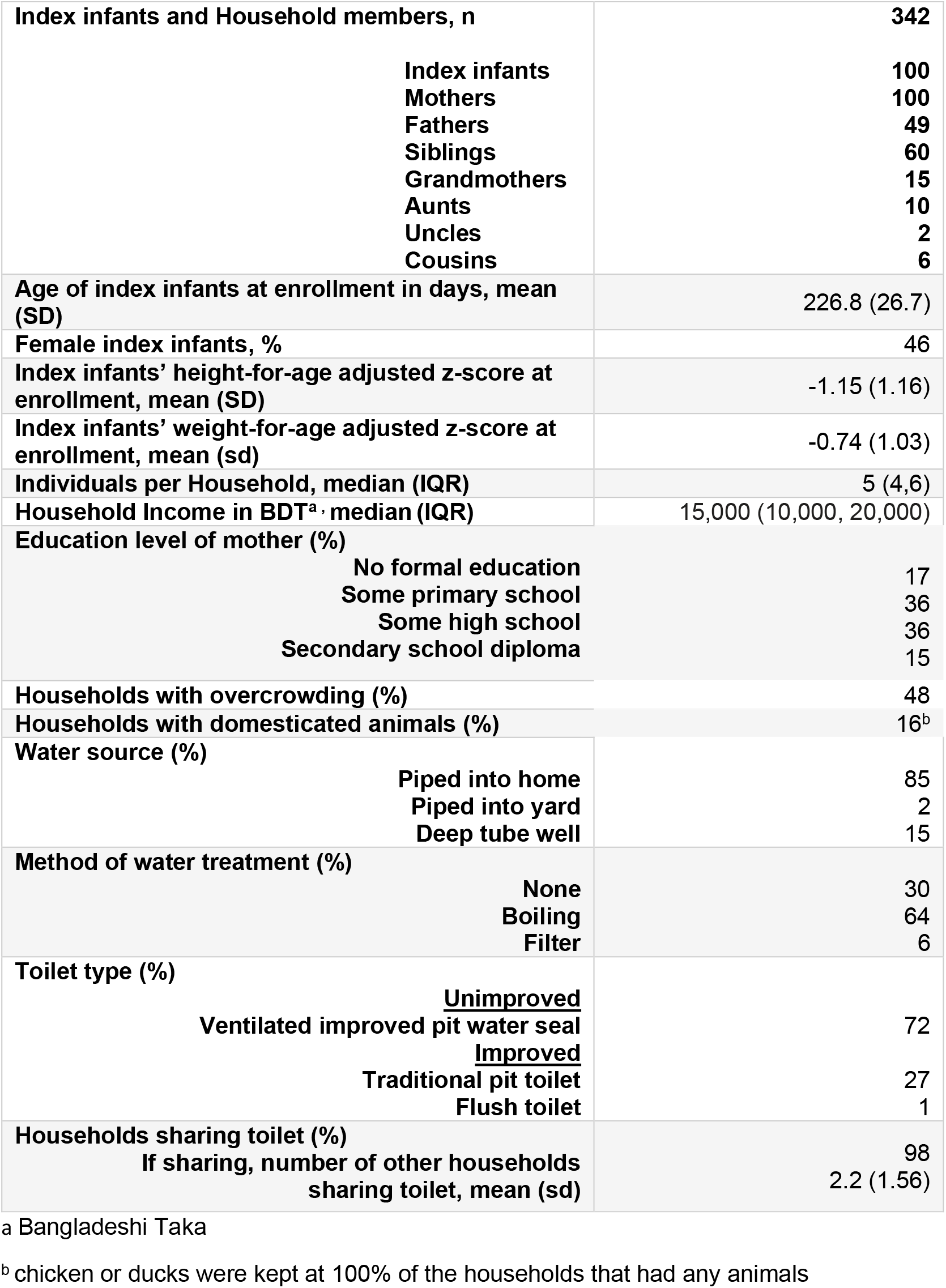
Characteristics of index infants and household members at enrollment.

In total, 9,774 surveillance stool samples were collected, with a 1.5% incidence of *Cryptosporidium spp* positivity and in the 384 diarrheal stool samples, Cryptosporidium positivity was 6.8%. Among index infants, *Cryptosporidium* positivity was 3.9% in surveillance stools and 7.6% in diarrheal stools (Table 2). Among the 100 index infants, there were 239 episodes of diarrhea. The mean duration of diarrhea was 7.6 days (sd 9.6) (Table 3). Index infants had more episodes of diarrhea and longer duration of diarrhea than all other age groups.

**Table 2.** *Cryptosporidium* positive diarrheal and surveillance stools collected during 8 months of follow-up, listed by age group.

|  | **Diarrheal stools,**<br>***Cryptosporidium***<br>**positive, n (%)** | **Surveillance**<br>**stools,**<br>***Cryptosporidium***<br>**positive, n (%)** | **Total stools,**<br>***Cryptosporidium***<br>**positive, n (%)** |
| --- | --- | --- | --- |
| Index children | 23 (7.6%) | 106 (3.9%) | 129 (4.2%) |
| Children, 1 to 5 years | 2 (20%) | 20 (2.8%) | 22 (3.0%) |
| Children, 5-10 years | 0 (0%) | 5 (0.5%) | 5 (0.5%) |
| Children, 10-17 years | 0 (0%) | 3 (0.8%) | 3 (0.8%) |
| Adults | 1 (2%) | 16 (0.3%) | 17 (0.3%) |
| All | 26 (6.8%) | 150 (1.5%) | 176 (1.7%) |

**Table 3.** Duration of diarrhea by age group

| **Age group (years)** | **N of episodes of**<br>**diarrhea** | **Mean duration in**<br>**days (sd)** |
| --- | --- | --- |
| < 1 | 239 | 7.6 (9.6) |
| 1-5 | 11 | 3.4 (2.2) |
| 6-10 | 13 | 5 (5.0) |
| 11-17 | 8 | 3 (1.2) |
| >=17 | 54 | 2.9 (4.4) |

When we considered *Cryptosporidium* infections per participant, children under age 5 carried the greatest burden of *Cryptosporidium* infection. The infected individuals included 44% of index infants, 35% of 2-5 years old, 15% of 6-10 years old, 20% of 11-17 year-olds with at least one *Cryptosporidium* infection during the 8-month follow up period. Among adult caregivers, 6% of mothers and 2% of fathers tested positive for *Cryptosporidium*. In total, 8 out of 170 (5%) adults tested positive for Cryptosporidium at least once during the study.

Shedding of *Cryptosporidium* occurred on average for 19.9 days (sd 8.3) among index infants (Table 4). Index infants had more prolonged shedding than other age groups (Figures 1 and 2). In the linear regression model, older children had shorter duration of shedding compared to index infants (beta -7.6; 95% CI -13, -2.4) (Supplemental Table 1). As expected, among index infants, we found that growth faltering (HAZ <=-2) was significantly associated with longer duration of shedding (beta 8.0; 95% CI 0.41, 16) (Supplemental Table 2). Presence of animals in the home and treatment of drinking water was negatively associated with duration of shedding in index children.

**Table 4.** Duration of *Cryptosporidium* shedding by age group.

| **Age group (years)** | **N of episodes of**<br>***Cryptosporidium***<br>**infection** | **Mean duration in**<br>**days (sd)** |
| --- | --- | --- |
| < 1 | 47 | 19.9 (8.3) |
| 1-5 | 10 | 16.1 (11.6) |
| 6-10 | 5 | 7.2 (1.1) |
| 11-17 | 3 | 7.0 (0) |
| >=17 | 8 | 16.2 (12.8) |

**Figure 1.**
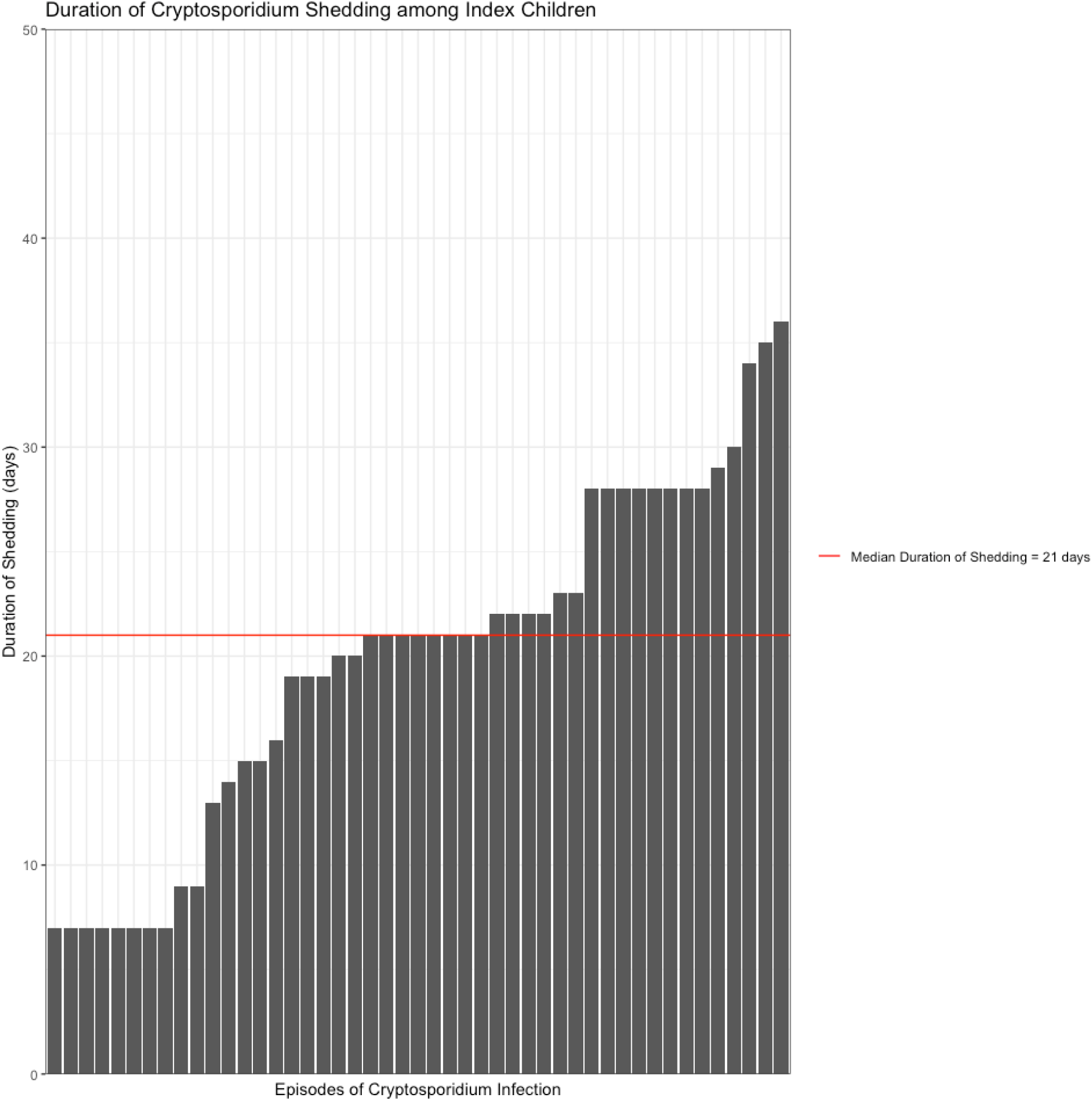
Duration of Cryptosporidium shedding by episode among index children. Each episode of Cryptosporidium infection captured is represented on the x-axis, and the duration in days is depicted on the y-axis (n = 47 episodes).

**Figure 2.**
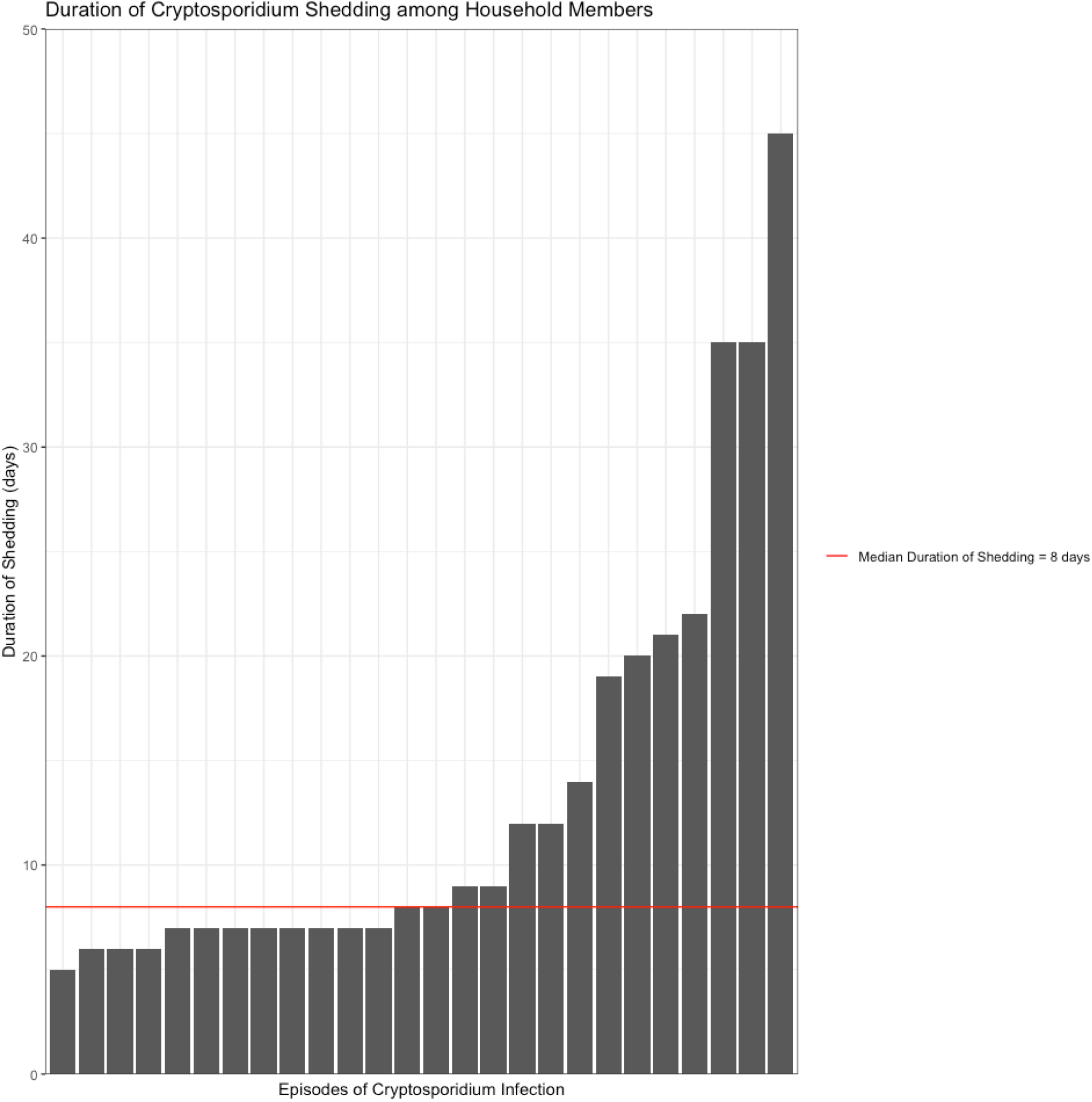
Duration of Cryptosporidium shedding among household members. Each episode of Cryptosporidium infection captured is represented on the x-axis, and the duration in days is depicted on the y-axis (n = 26 episodes).

Repeat infections only occurred in young children. Three index infants, plus one unrelated sister, experienced repeat infections, and all were age three years and younger (Table 5). Interestingly, three of the four children had a longer duration of parasite shedding during their second infection compared to their first infection.

**Table 5.** Duration of *Cryptosporidium* shedding in individuals with repeat infections.

| Participant | Age (years) | Sex | Duration of shedding in 1<sup>st</sup> episode (days) | Duration of Shedding in 2<sup>nd</sup> episode (days) |
| --- | --- | --- | --- | --- |
| Index Child | <1 | Female | 7 | 7 |
| Index Child | <1 | Male | 7 | 29 |
| Index Child | <1 | Female | 21 | 28 |
| Sister | 3 | Female | 7 | 20 |

Figure 3 shows weekly stool results among all 44 children who tested positive for *Cryptosporidium* in at least one weekly sample over the 8-month follow up period. We observed that not all diarrheal episodes were positive for *Cryptosporidium*, and in some cases a child may have had multiple diarrheal episodes and only a single weekly positive *Cryptosporidium* infection. And yet others had prolonged shedding spanning several weeks but none or limited diarrhea.

**Figure 3.**
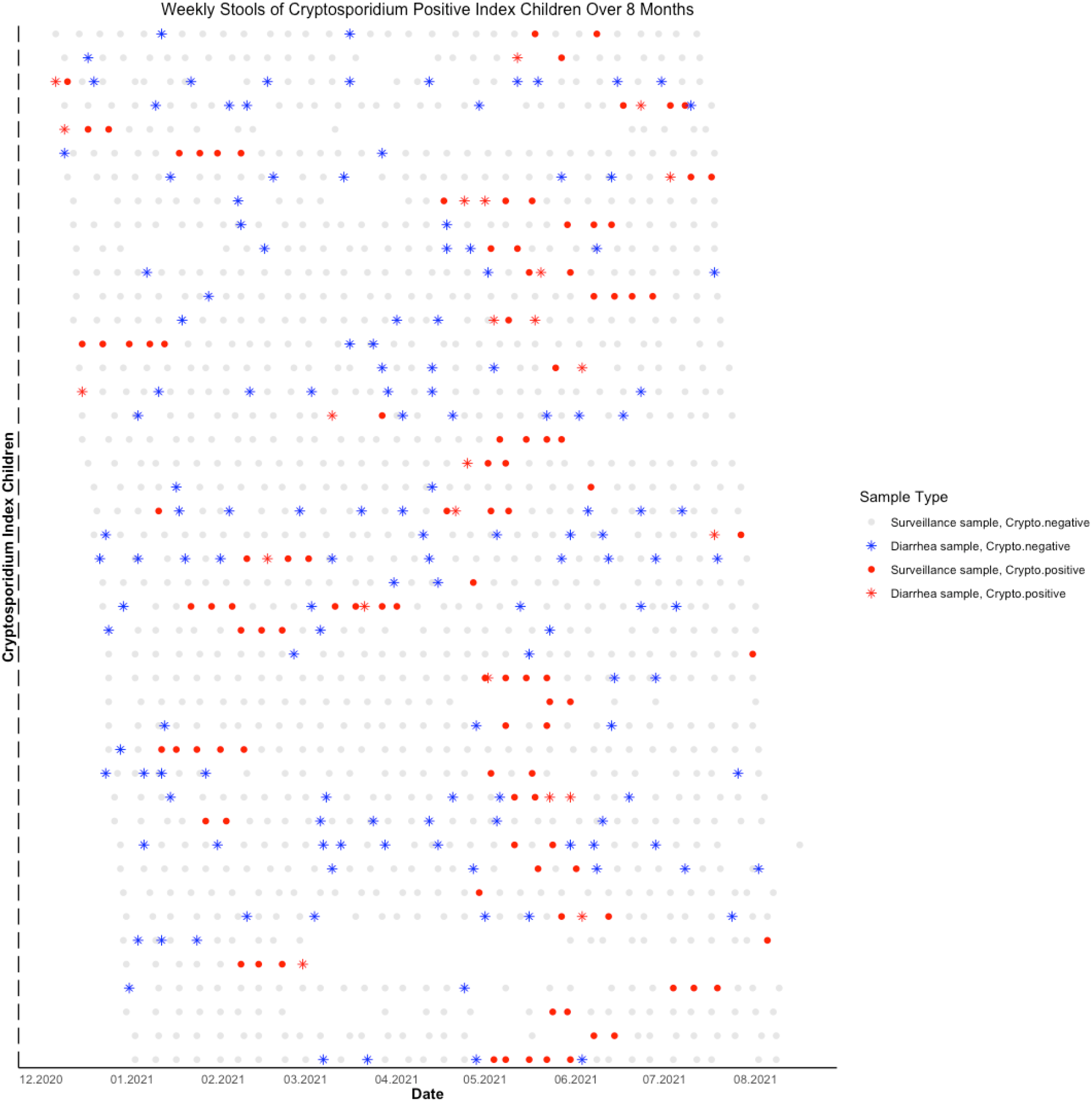
*Cryptosporidium* shedding in index children. This figure represents all weekly stools from all index children who had at least one positive *Cryptosporidium* sample over the 8-month follow up period. Cryptosporidium positive stools are shaded in red, and a diarrheal stool is represented by (*). Areas that are blank represent missing stool samples. Date of sample collection is listed on the x-axis.

The monthly rate of *Cryptosporidium* positivity among stool samples was consistent throughout the study, however, there was a large increase in infections during May 2021 (Figure 4) [20]. There was a spike in COVID-19 cases in April 2020, after which the Bangladesh government mandated a strict lockdown. These restrictions were lifted in May 2020, which is when the corresponding spike in *Cryptosporidium* cases is observed [21]. Of all the enrolled participants (n=342), 30.4% were SARS-CoV-2 IgG positive at baseline, and this percent increased to 45.8% at the end of the study. Eighty-nine participants went from a negative to positive SARS-CoV-2 serology during the 8-month follow up period. SARS-CoV-2 IgG positivity in children was not associated with markers of malnutrition, there were no observed differences in positivity by height-for-age or height-for-weight adjusted z-scores (t-test, p> 0.05).

**Figure 4.**
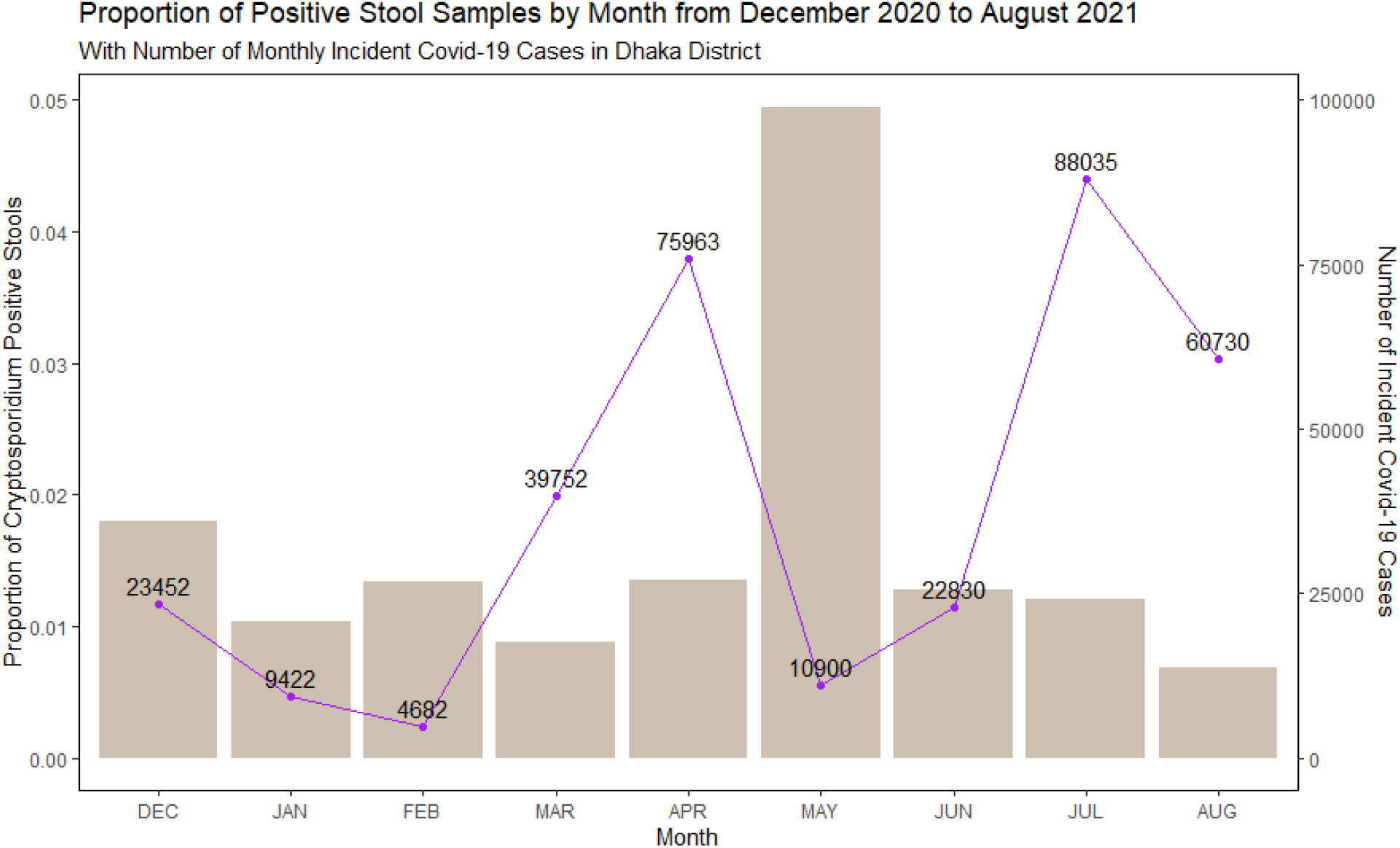
*Cryptosporidium* positive stool samples by month. Rate of Cryptosporidium positivity was stable until May 2021, when there was a large spike in Cryptosporidium cases. The total number of local COVID-19 cases are overlaid and demonstrate a spike in April 2021.

## DISCUSSION

In this prospective, longitudinal study of infants and families living in an urban area in Bangladesh, there was a high rate of *Cryptosporidium* infection and shedding in young children with or without diarrhea. We also observed that household family members had higher rates of infection including 35% percent of children 2-5 years and 6% of mothers.

Shedding of *Cryptosporidium* lasted from 5 to 36 days among index infants. Although infection rates were lower in older ages, prolonged shedding was observed across the age spectrum. Notably, even in adult participants, we found *Cryptosporidium* shedding to range from 5 to 35 days. We observed *Cryptosporidium* shedding in both diarrheal and non-diarrheal infections. Interestingly, growth faltering was associated with prolonged duration of shedding. Presence of domestic fowl in the home and treatment of drinking water were marginally inversely associated with duration of shedding.

Treatment of drinking water may be directly related to decreased shedding by reducing the burden of parasite exposure through participants’ gastrointestinal tracts. Alternatively, treatment of drinking water may be a proxy for other conditions that predispose to reduced parasite shedding.

The only other study that has looked at *Cryptosporidium* shedding with the same intensity as the present study was in Scotland in 1988 looking at immunocompetent children and adults ages 9 months to 88 years. The Scottish study also reported a range of *Cryptosporidium* shedding from 2 -35 days [22] using daily household follow up of all *Cryptosporidium* positive patients from their region. This differs from modeling-based projections which have reported longer shedding times but may represent a lack of precision in those models [23].

We did not observe any reduced duration in shedding in second versus primary infection, suggesting that prior infection does not protect or prime for repeat episodes of shedding. The duration of oocyst shedding is important for transmission, as it represents the period of time an individual is infectious and can spread the infection to close contacts. The prolonged shedding observed in all age groups provides multiple opportunities for transmission within a household. In addition, we report shedding not only in the diarrheal episodes but also in surveillance samples. Efforts to treat or disrupt *Cryptosporidium* infections that solely target diarrheal infections will not be adequate to control transmission.

Our data, combined with the Scottish data, demonstrate that in a healthy population or in a malnourished population, *Cryptosporidium* shedding still occurs for a prolonged period, especially in children. Interventions to prevent *Cryptosporidium* infection in vulnerable populations must address the fact that shedding is occurring even when participants are not experiencing diarrhea. With this prolonged shedding in the household environment, and the failure of WASH in other studies, pharmacologic treatment of non-diarrheal infection in children and perhaps entire households may be needed, as this may be the only way to eliminate *Cryptosporidium* from the household environment.

This study is unique in that it took place during the SARS-CoV-2 pandemic. Nearly 1/3^rd^ of participants had a positive SARS-CoV-2 IgG response. After enacting a series of nationwide lockdowns in 2020, all restrictions on movement of individuals were lifted in September 2020. Seven months later, Bangladesh experienced a record number of COVID-19 cases in April 2021, and the government imposed a strict lockdown once again. After the spike in COVID-19 cases in April 2021, we observed a corresponding spike in *Cryptosporidium* cases in May 2021 [20]. We propose the corresponding spike in *Cryptosporidium* cases could be attributed to the nationwide lockdown, as all family members were required to remain within their homes. Therefore, while social distancing was enacted to protect against spread of SARS-CoV-2, it may have inadvertently reduced social distancing in the home, allowing for increased *Cryptosporidium* transmission among household contacts. One limitation is the SARS-CoV-2 data is at a country-wide level. If we had accurate rates of SARS-CoV-2 infection in the Mirpur region, it would have been possible to link social distancing measures for SARS-COV-2 to *Cryptosporidium* cases at a community level.

This is the largest study to follow entire households over 8 months to describe incidence of cryptosporidiosis in an endemic region. Notably, we found that prolonged shedding of cryptosporidiosis occurs among infants and adults, increasing the risk for exposure. Determining methods for interrupting transmission within the household should be prioritized to protect infants from cryptosporidiosis, especially given the lack of vaccine and approved-pharmacologic treatments in this vulnerable population.

## Data Availability

All data produced in the present study will be made available upon reasonable request to the authors.

## ACKNOWLEDGEMENTS

We thank the families of Mirpur for participating in this study.

## SUPPLEMENTAL TABLES

**Supplemental Table 1.** Multiple linear regression to calculate duration of *Cryptosporidium* shedding based on age among all participants.

| Characteristic | Change in shedding duration (days) | 95% CI<sup>1</sup> | p-value | GVIF<sup>1</sup> | Adjusted GVIF<sup>12</sup> |
| --- | --- | --- | --- | --- | --- |
| **(Intercept)** | 21 | 17, 24 | **<0.001** |  |  |
| **Age category** |  |  | **0.017** | 1.1 | 1.0 |
| *Index infants* | — | — |  |  |  |
| *1 -17 years* | -7.6 | -13, -2.4 | **0.005** |  |  |
| *17-65 years* | -2.7 | -10, 4.5 | 0.5 |  |  |
| **Sex** | -0.78 | -5.3, 3.7 | 0.7 | 1.1 | 1.0 |
| **Positive COVID-19 serology** | -2.2 | -7.6, 3.2 | 0.4 | 1.1 | 1.1 |

<sup>1</sup>CI = Confidence Interval, GVIF = Generalized Variance Inflation Factor
<sup>2</sup>GVIF<sup>1</sup>^(1/(2\*df))

**Supplemental Table 2.**
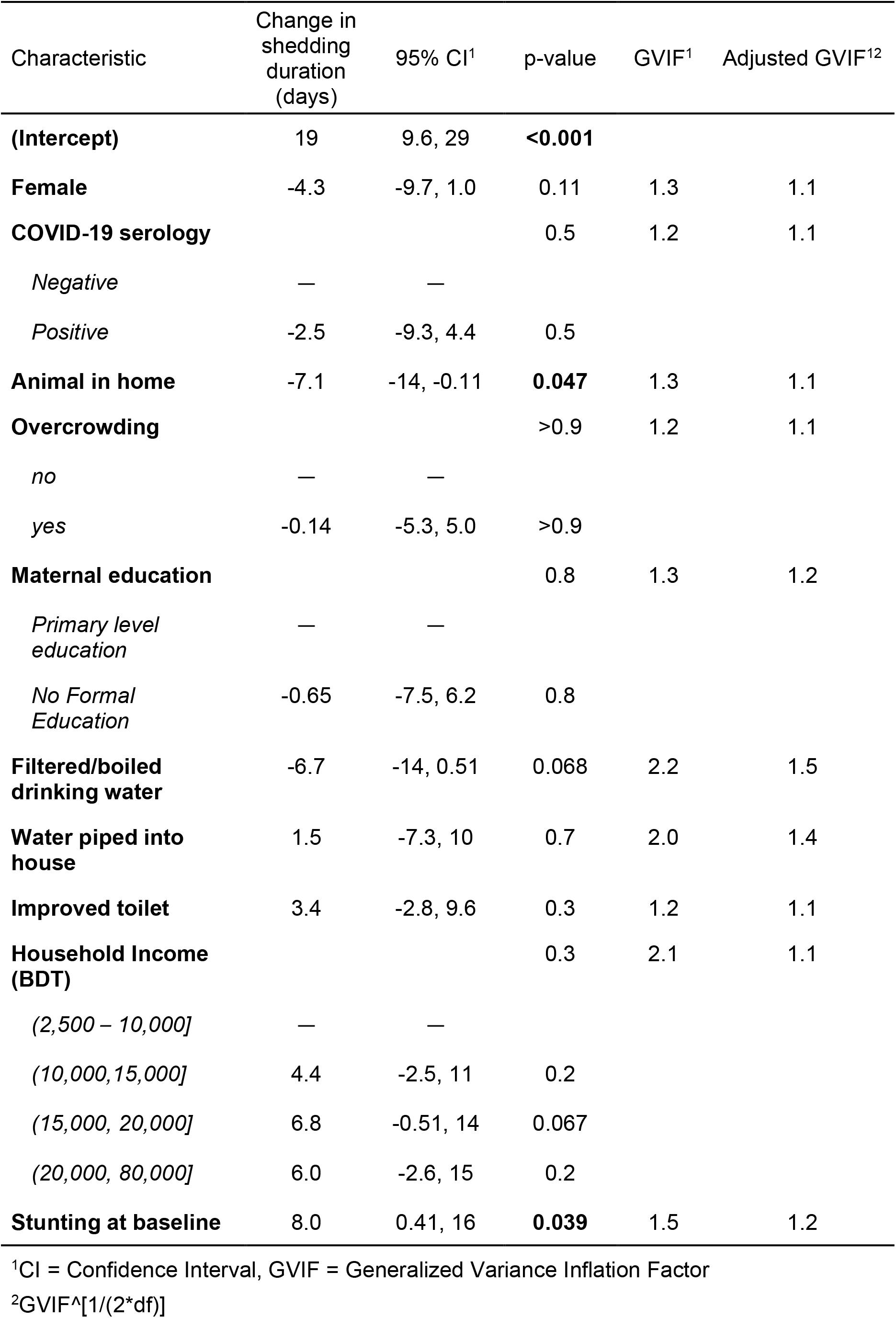
Multiple linear regression to calculate duration of Cryptosporidium shedding based on risk factors among index infants

